# Home-Based Transcutaneous Auricular Vagus Nerve Stimulation for Generalized Anxiety Disorder: Safety, Feasibility, and Preliminary Clinical Outcomes in a Single-Arm Prospective Study

**DOI:** 10.64898/2026.06.02.26354707

**Authors:** Mohsen Mosayebi Samani, Erfan Zahirmardi, Saman Hedayat fard, Serjhik Azerians

**Affiliations:** Department of Psychology and Neuroscience, Leibniz Research Center for Working Environment and Human Factors, Dortmund, Germany; Department of Psychiatry, Ibn Sina Psychiatric Hospital, Bandar Abbas, Iran

## Abstract

**Background:** Generalized anxiety disorder (GAD) is associated with substantial psychological burden, autonomic dysregulation, and limitations of existing pharmacological and psychotherapeutic treatments. Transcutaneous auricular vagus nerve stimulation (taVNS) has emerged as a promising non-invasive neuromodulation approach, but evidence regarding home-based application in GAD remains limited.

**Objective:** To evaluate the feasibility, safety, and preliminary clinical and physiological outcomes of a home-based taVNS intervention in adults with psychologist-confirmed moderate-to-severe GAD.

**Methods:** In this prospective single-arm feasibility study, 48 participants initiated a 4-week home-based taVNS intervention consisting of two daily stimulation sessions performed five days per week. Clinical assessments were conducted at baseline, Week 2, Week 4, and follow-up visits at Weeks 6 and 8. Ambulatory electrocardiographic monitoring was performed before treatment initiation, at Week 2, and at the end of treatment to assess heart rate variability (HRV) using the root mean square of successive differences (RMSSD). Primary outcomes included feasibility, safety, adherence, and change in clinician-rated anxiety severity (HAM-A).

**Results:** Thirty-four participants completed the study and were included in the primary analyses. HAM-A scores decreased significantly from baseline to Week 4 (estimated mean difference [EMD] −6.9, 95% CI −10.4 to −3.4, p = 0.001), with partial maintenance during follow-up. Improvements were also observed in Beck Anxiety Inventory scores, whereas changes in GAD-7, perceived stress, depressive symptoms, and sleep quality were not statistically significant. RMSSD increased significantly from baseline to Week 4 (EMD 6.7 ms, 95% CI 2.1–11.3, p = 0.009). Greater increases in RMSSD were associated with larger reductions in HAM-A (R² = 0.18, p = 0.031) and BAI scores (R² = 0.21, p = 0.019). No serious adverse events occurred. Mean adherence was 79.8%, and 73.5% of participants completed at least 70% of prescribed stimulation sessions.

**Conclusions:** Home-based taVNS was feasible and generally well tolerated in adults with moderate-to-severe GAD. Preliminary improvements in clinician-rated anxiety severity and autonomic physiological measures were observed; however, the single-arm design precludes causal inference. These findings support further evaluation of home-based taVNS in adequately powered randomized sham-controlled trials.

## 1. Introduction

Anxiety disorders are among the most prevalent psychiatric disorders worldwide and represent a major contributor to disability, healthcare utilization, reduced occupational functioning, and impaired quality of life (Alonso et al., 2018; Bandelow and Michaelis, 2015) . Among these conditions, Generalized Anxiety Disorder (GAD) is characterized by excessive and difficult-to-control worry occurring on most days for at least several months, accompanied by autonomic hyperarousal, muscular tension, irritability, impaired concentration, restlessness, and sleep disturbance, and is associated with substantial psychosocial burden, increased cardiovascular morbidity, impaired daily functioning, and elevated long-term healthcare costs ((DeMartini et al., 2019; Stein Murray and Sareen, 2015).

Despite the availability of pharmacological and psychotherapeutic interventions, treatment accessibility and long-term effectiveness remain limited (Alonso et al., 2018; Cuijpers et al., 2024; Leichsenring et al., 2022). Cognitive behavioral therapy (CBT) demonstrates robust efficacy but is frequently constrained by therapist availability, long waiting lists, and cost (Levy et al., 2021; Loerinc et al., 2015).

Pharmacological treatments including selective serotonin reuptake inhibitors (SSRIs) and serotonin-norepinephrine reuptake inhibitors (SNRIs) provide moderate symptom improvement but are commonly associated with adverse effects, incomplete remission, relapse after discontinuation, and poor long-term adherence (Halstead et al., 2025; Reinhold and Rickels, 2015; Roest et al., 2015). Together, these limitations highlight the need for alternative intervention strategies capable of addressing both the psychological and physiological dimensions of GAD.

Increasing evidence suggests that mental components of GAD such as excessive worry and fear implicate disrupted functional connectivity across prefrontal–limbic circuits involving the amygdala, anterior cingulate cortex, insula, hippocampus, and locus coeruleus, regions critically involved in emotional regulation and autonomic control (Hilbert et al., 2019; Shin and Liberzon, 2010). In parallel, physiological studies indicate that autonomic dysregulation represents an important neurophysiological component of GAD, reflected by reduced vagal tone, elevated sympathetic activation, altered heart rate variability (HRV), and dysregulated stress-axis function (Bandelow et al., 2017; Chalmers et al., 2014; Friedman, 2007). In addition, the somatic symptoms frequently observed in GAD, including muscular tension, restlessness, trembling, sweating, dizziness, gastrointestinal discomfort, and fatigue, are thought to reflect altered interoceptive processing and heightened bodily threat perception involving the insula and anterior cingulate cortex, regions implicated in the integration of emotional and autonomic states (Craig, 2009; Hilbert et al., 2014; Paulus and Stein, 2010, 2006; Stein and Craske, 2017). Together, these findings support the growing interest in neuromodulatory interventions aimed at restoring prefrontal–limbic network balance and autonomic regulation.

Transcutaneous auricular vagus nerve stimulation (taVNS) has emerged as a promising and safe neuromodulatory approach capable of modulating these networks through non-invasive stimulation of the auricular branch of the vagus nerve (Colzato and Beste, 2020; Farmer et al., 2021; Kim et al., 2022). Neurophysiological and neuroimaging studies including paired-pulse transcranial magnetic stimulation, electroencephalography (EEG), and functional magnetic resonance imaging (fMRI) showed that taVNS can modulate activity in the nucleus tractus solitarius and downstream vagal projection regions, including brainstem, limbic, thalamic, insular, and prefrontal networks (Boscarol et al., 2025; Fallgatter et al., 2003; Fang et al., 2016; Frangos et al., 2015; Yakunina et al., 2017). In addition, physiological studies suggest that non-invasive vagus nerve stimulation can reduce sympathetic nerve activity and modulate HRV-related autonomic indices, supporting its relevance for interventions targeting autonomic imbalance (Badran et al., 2018; Clancy et al., 2014; Machetanz et al., 2021). Conflicting results have however also been reported, likely due to heterogeneity in stimulation parameters, target locations, recording methods, and participant characteristics, highlighting the need for further systematic evaluation (Keute et al., 2021; Wolf et al., 2021).

Previous taVNS studies have reported beneficial effects across neurological and psychiatric disorders including epilepsy, insomnia, and major depressive disorder (Bauer et al., 2016; Hein et al., 2013; Lampros et al., 2021; Rong et al., 2016; Zhang et al., 2024). Anxiety-relevant evidence is also emerging. In one study, tVNS was reported to facilitate fear extinction and reduce psychophysiological indices of stimulus-specific fear in experimental and phobic populations (Szeska et al., 2025, 2020). Additional studies suggest that tVNS may reduce stress-related sympathetic responses in PTSD and improve anxiety symptoms in Parkinson’s disease with comorbid anxiety (Gurel et al., 2020; Wang et al., 2026). Thus, despite mechanistic plausibility and emerging anxiety-relevant evidence, the direct effects of home-based taVNS in adults with primary GAD remain insufficiently evaluated.

The present prospective single-arm observational study therefore aimed to evaluate the feasibility, safety, and preliminary clinical effects of a home-based taVNS system in adults with psychologist-confirmed GAD. Participants completed taVNS twice daily for four weeks followed by post-intervention assessments at two weeks and one month after stimulation discontinuation. In addition, longitudinal physiological monitoring was performed to evaluate exploratory autonomic physiological changes associated with symptom improvement. We hypothesized that: (1) participants would demonstrate reductions in clinician-rated anxiety severity following four weeks of intervention; (2) home-based taVNS would demonstrate acceptable safety, and adherence; and (3) anxiety improvement would be accompanied by favorable modulation of autonomic physiological biomarkers including HRV.

## 2. Methods

### 2.1 Study Design

This study was designed as a prospective single-arm feasibility study investigating the safety and preliminary clinical effects of home-based taVNS in adults with psychologist-confirmed moderate-to-severe GAD. Secondary objectives were to evaluate adherence and exploratory physiological changes associated with clinical improvement. The study consisted of a baseline clinical assessment, a four-week home-use taVNS intervention period consisting of twice-daily stimulation sessions performed five days per week, and post-intervention follow-up assessments at two weeks and one month after stimulation discontinuation. Psychological assessments were performed at baseline, Week 2, Week 4, two-week and one-month follow-up. Physiological monitoring was conducted longitudinally during three predefined windows: the first days before stimulation initiation as the individual physiological baseline (about 8 hours of analyzable physiological recording), around Week 2 to capture mid-intervention physiological change, and one day after Week 4 to assess end-of-treatment physiological status. This repeated-window approach was selected to reduce participant burden while still allowing within-participant characterization of physiological trajectories across the intervention, Figure 1.

**Figure 1.**
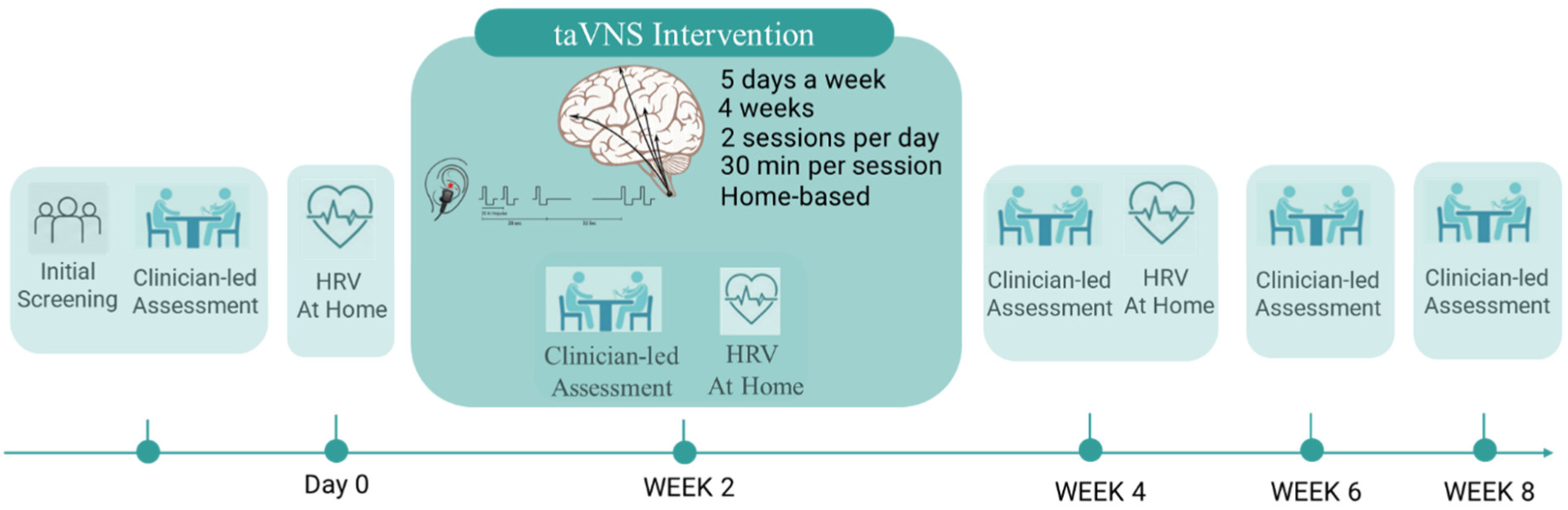
Study design and assessment schedule. Participants underwent screening and baseline evaluation prior to initiating a 4-week home-based transcutaneous auricular vagus nerve stimulation (taVNS) intervention. Clinical assessments were conducted at baseline, Week 2, Week 4 (end of treatment), and follow-up visits at Weeks 6 and 8. Home-based HRV monitoring was performed before treatment initiation, at Week 2, and at the end of treatment. The intervention consisted of two 30-min stimulation sessions per day, five days per week, for four weeks.

The study protocol was approved by the local ethics committee (Hormozgan University of Medical Sciences Ethical Committee; IR.HOMS.REC.1402.123). All participants received written and verbal information about study procedures, stimulation, wearable monitoring, data handling, and their right to withdraw without consequences for clinical care. Written informed consent was obtained before any study-specific assessment or physiological recording.

### 2.2 Participants

Participants aged 18–65 years were recruited from outpatient CBT waiting lists. Potentially eligible patients were contacted during the CBT waiting-list period and informed about the study. After written informed consent, participants completed standard self-reported questionnaires and underwent a repeat DSM-5-oriented psychologist-led clinical assessment supported by Hamilton Anxiety Rating Scale (HAM-A) scoring. This psychologist-led assessment served as the clinical reference assessment for diagnosis, eligibility confirmation, and interpretation of symptom change across the study.

A HAM-A score of at least 25 was required to ensure inclusion of clinically meaningful moderate-to-severe GAD. In addition, self-reported questionnaires (see section 2.3) were recorded prior to the psychologist assessment to characterize anxiety, somatic anxiety, perceived stress, depressive symptoms, and sleep quality, but they were not used as independent diagnostic reference standards.

Participants were excluded if the psychologist-led assessment identified a dominant psychiatric or medical condition likely to confound physiological interpretation or clinical outcome assessment. These conditions included primary major depressive episode, post-traumatic stress disorder, bipolar disorder, psychotic disorder, panic disorder as the dominant diagnosis, insomnia disorder as the dominant clinical condition, active substance dependence, severe cardiovascular disease, neurological disorders, pregnancy, beta-blocker use, implanted electrical medical devices, current neuromodulation therapy, or insufficient wearable recording quality. Participants were also excluded if they changed psychotropic medication or initiated CBT between the repeat clinical assessment and completion of wearable monitoring. Stable SSRI/SNRI use was permitted only if dosage had remained unchanged for at least eight weeks before monitoring. Menstrual phase and hormonal contraceptive use were recorded because of their potential influence on autonomic physiological trajectories across the intervention.

### 2.3 Psychological measures

The HAM-A is a clinician-administered instrument assessing both psychic and somatic components of anxiety across 14 symptom domains, including anxious mood, tension, fears, insomnia, cardiovascular symptoms, gastrointestinal symptoms, and autonomic complaints. Each item is rated on a 5-point scale ranging from 0 (“not present”) to 4 (“severe”), resulting in total scores ranging from 0 to 56, with higher scores indicating greater anxiety severity (Hamilton, 1960; Maier et al., 1988). Conventional interpretation thresholds categorize scores below 17 as mild anxiety, scores between 18 and 24 as mild-to-moderate anxiety, and scores between 25 and 30 as moderate-to-severe anxiety severity (Matza et al., Int J Methods Psychiatr Res, 2010). The HAM-A was administered by a certified clinician at baseline, Week 2, Week 4 (end of stimulation), two-week and one-month follow-up. Clinical response was operationally defined as a reduction of at least 50% from baseline score. Remission was defined as a HAM-A total score of 7 or lower, consistent with previous anxiety-treatment studies (Baldwin et al., 2011; Bandelow et al., 2017). The GAD-7 is a seven-item self-report scale assessing core generalized anxiety symptoms over the previous two weeks. Each item is scored from 0 (“not at all”) to 3 (“nearly every day”), yielding total scores ranging from 0 to 21, with higher scores indicating greater symptom severity. Conventional cut-offs of 5, 10, and 15 represent mild, moderate, and severe anxiety symptoms, respectively (Spitzer et al., n.d.)

The BAI is a 21-item self-report questionnaire emphasizing somatic and physiological symptoms of anxiety, including numbness, trembling, dizziness, sweating, heart pounding, and fear of losing control. Each item is rated on a 4-point scale from 0 (“not at all”) to 3 (“severely — I could barely stand it”), resulting in total scores ranging from 0 to 63. Standard interpretation thresholds categorize scores of 0–7 as minimal anxiety, 8–15 as mild anxiety, 16–25 as moderate anxiety, and 26–63 as severe anxiety ((Beck et al., 1988).

Perceived stress was assessed using the PSS-10, a 10-item self-report questionnaire evaluating the degree to which situations in one’s life are appraised as stressful during the previous month. Each item is rated on a 5-point scale from 0 (“never”) to 4 (“very often”), yielding total scores ranging from 0 to 40, with higher scores indicating greater perceived stress. Commonly used interpretation ranges classify scores of 0–13 as low stress, 14–26 as moderate stress, and 27–40 as high perceived stress (Solis et al., 1983).

Depressive symptoms were assessed using the PHQ-9, a nine-item self-report scale evaluating depressive symptoms over the previous two weeks. Each item is scored from 0 (“not at all”) to 3 (“nearly every day”), resulting in total scores ranging from 0 to 27, with higher scores indicating greater depressive symptom severity. Conventional cut-offs of 5, 10, 15, and 20 indicate mild, moderate, moderately severe, and severe depressive symptoms, respectively (Kroenke et al., n.d.).

Sleep quality was measured using the PSQI, a self-report instrument assessing sleep quality and disturbances over the previous month across seven domains, including sleep duration, latency, efficiency, disturbances, and daytime dysfunction. Component scores are summed to generate a global score ranging from 0 to 21, with higher scores indicating poorer sleep quality. A global PSQI score greater than 5 is conventionally interpreted as indicating poor sleep quality (Buysse Charles F Reynolds Ill et al., n.d.).

### 2.4 Wearable Physiological Measures

Electrocardiogram (ECG) signals were acquired using the Movesense MD sensor (CE-certified medical device) at a sampling rate of 512 Hz ((Rogers et al., 2022). ECG was recorded from one bipolar channel using pre-gelled Ag/AgCl solid-gel electrodes placed on the thorax in a chest-lead configuration suitable for robust R-wave detection and heart rate variability analysis. Electrode sites were selected to maximize QRS amplitude and reduce motion-related artifacts while maintaining stable electrode–skin contact throughout the recording period. The skin was prepared before electrode attachment, and electrodes with stable gel contact were used to reduce impedance and improve signal quality.

Raw ECG data were processed offline using Kubios HRV Scientific software (University of Eastern Finland, Kuopio, Finland). ECG preprocessing included R-peak detection, derivation of beat-to-beat R–R interval sequences, detrending, and automatic artifact correction according to the robust beat classification algorithm described by Lipponen and Tarvainen (Lipponen and Tarvainen, 2019). Heart rate variability (HRV) was quantified using the root mean square of successive differences (RMSSD), calculated as the square root of the mean squared differences between successive R–R intervals, reflecting short-term parasympathetic cardiac modulation.

### 2.5 Transcutaneous Auricular Vagus Nerve Stimulation

Stimulation was delivered using the medically certified AXION STIM-PRO X9+ device (AXION GmbH, Germany) with a research-grade auricular electrode positioned at the cymba conchae, targeting the auricular branch of the vagus nerve. Prior to each stimulation session, participants applied conductive Ten20 paste to the electrode–skin interface to reduce tissue–electrode impedance and improve stimulation consistency. taVNS was applied using biphasic charge-balanced pulses with a pulse width of 250 microseconds. Stimulation was delivered in bursts at 25 Hz during a 28-second ON / 30-second OFF duty cycle for a total session duration of 30 minutes. Stimulation intensity was individually adjusted according to participant sensory perception and tolerability, up to a maximum output intensity of 5 mA within predefined safety limits. Participants performed stimulation sessions twice daily, five days per week, for four consecutive weeks under home-use conditions.

### 2.6 Safety, Adverse Events, and Adherence

Safety and adverse effects were evaluated throughout the intervention and follow-up using structured adverse-event questioning (Farmer et al., 2021; Kim et al., 2022; Redgrave et al., 2018). Adverse events were classified by severity, duration, outcome, and relationship to the device. Events of interest included local skin irritation, pain or discomfort at the stimulation site, headache, dizziness, nausea, sleep disturbance, and any cardiovascular or neurological symptoms. Serious adverse events were defined as events resulting in hospitalization, life-threatening condition, persistent disability, or death. Stimulation adherence was quantified using the percentage of prescribed sessions completed with per-protocol adherence predefined as completion of at least 70% of prescribed stimulation sessions, consistent with commonly used adherence thresholds in behavioral and neuromodulation studies (Jin et al., 2020).

### 2.7 Experimental Procedure

Following initial screening and written informed consent, participants first completed baseline self-reported questionnaires including the GAD-7, BAI, PSS-10, PHQ-9, and PSQI and underwent subsequently a DSM-5-oriented psychologist-led clinical assessment performed by a trained clinician to confirm eligibility according to the predefined study criteria. Following confirmation, participants were onboarded to the stimulation and physiological monitoring systems and received standardized training regarding stimulation-device handling, electrode positioning, ECG sensor placement, physiological recording procedures, safety precautions, and troubleshooting, Figure 1.

During the first home-monitoring day before stimulation initiation, participants recorded a about of 8 hours of physiological measurements under naturalistic conditions to establish the individual baseline physiological profile. Participants were instructed to maintain their usual daily routines during recording periods. Afterwards, participants initiated self-administered home-based taVNS beginning the following day. Participants performed taVNS sessions twice daily for five days per week over four consecutive weeks. Mid-intervention physiological monitoring was performed around Week 2 while participants continued stimulation according to protocol. End-of-intervention physiological monitoring was performed after completion of the Week 4 intervention period. Clinical and psychological reassessments were performed at Week 2 and Week 4. Following completion of the stimulation phase, participants entered a stimulation-free follow-up period with assessments at two weeks and one month after stimulation discontinuation to evaluate persistence of clinical effects, early relapse, and delayed adverse events, Figure 1.

Participants were instructed to contact the study team in the event of discomfort, adverse symptoms, technical problems, or uncertainty regarding stimulation procedures. Compliance, stimulation tolerability, and recording quality were reviewed regularly throughout the study period

### 2.8 Statistical Analysis

Data analysis was conducted using Python 3.11 with the SciPy and Statsmodels libraries.

#### 2.8.1. Psychological Outcome Analysis

Psychological outcomes were analyzed across five predefined time points: baseline (BL), Week 2, Week 4 (end of stimulation), two-week post-intervention follow-up (Week 6), and one-month post-intervention follow-up (Week 8), using linear mixed-effects modeling. Time was modeled as a categorical fixed effect, while participant-specific random intercepts were included to account for repeated within-subject measurements and inter-individual baseline variability. The overall effect of time was first evaluated using omnibus F-tests derived from the mixed-effects models. If the overall model was significant, post hoc contrasts comparing each follow-up time point against baseline were subsequently evaluated and corrected using the Holm multiple-comparison procedure. Estimated mean differences (EMD), 95% confidence intervals (CI), model-derived p values, and standardized within-subject effect sizes were reported to characterize the magnitude and temporal pattern of change.

The **primary** analysis evaluated within-participant longitudinal change in clinician-rated anxiety severity using the HAM-A. The primary clinical endpoint was change in HAM-A score from baseline to Week 4. **Secondary** psychological outcomes included longitudinal changes in GAD-7, BAI, PSS-10, PHQ-9, and PSQI scores across the same five assessment time points.

Clinical response was operationally defined as a reduction of at least 50% in HAM-A score relative to baseline. Remission was defined as a HAM-A total score of 7 or lower at Week 4 or during follow-up assessments. Response and remission rates were summarized descriptively together with exact binomial 95% confidence intervals. Exploratory relapse analyses were additionally performed among Week 4 responders and defined as loss of response status during the stimulation-free follow-up period.

#### 2.8.2. Physiological Outcome Analysis

Physiological recordings were collected during daytime ambulatory conditions and processed following artifact rejection and signal-quality filtering. For each participant and assessment window, RMSSD values were averaged across the valid recording period to obtain a single representative estimate of parasympathetic autonomic activity, consistent with established heart rate variability methodological recommendations for long-duration recordings (Laborde et al., 2017). RMSSD was analyzed across the predefined physiological monitoring windows using repeated-measures linear mixed-effects models with monitoring window as a categorical fixed effect and participant-specific random intercepts. Exploratory models additionally evaluated the potential influence of age, sex, baseline anxiety severity, and stable SSRI/SNRI use as covariates where appropriate.

#### 2.8.3. Clinical–Physiological Association Analysis

Exploratory clinical–physiological association analyses were conducted to examine whether autonomic changes during treatment were associated with clinical improvement. Change scores from baseline to Week 4 were calculated for RMSSD and each psychological outcome measure. Separate linear regression models were performed using change in HAM-A (ΔHAM-A), BAI (ΔBAI), and GAD-7 (ΔGAD-7) scores as dependent variables and change in RMSSD (ΔRMSSD) as the independent variable. Regression coefficients (β), 95% confidence intervals, p values, and explained variance (R²) were reported for each model. These analyses were exploratory and the study was not powered for biomarker validation. Therefore, the findings were interpreted as hypothesis-generating and intended to inform future controlled investigations.

## 3. Results

### 3.1. Participant Characteristics and Study Flow

A total of 82 individuals were initially screened during the CBT waiting-list recruitment period. Of these, 61 individuals provided informed consent and agreed to participate in the study procedures. Following repeat psychologist-led diagnostic assessment and eligibility screening, 48 participants met all inclusion criteria and initiated the study intervention. Fourteen participants discontinued participation before study completion. Reasons for dropout included scheduling and adherence difficulties (n = 4), local stimulation discomfort (n = 5), and withdrawal because of perceived lack of benefit during the intervention period (n = 5). The final analytic sample therefore consisted of **34 participants** (38.7 ± 11.2; 21 Female) who completed baseline assessment and sufficient longitudinal clinical and physiological measurements for inclusion in the primary analyses, Table 1.

**Table 1.** Baseline Demographic and Clinical Characteristics (N = 34)

| **Table 1. Baseline Demographic and Clinical Characteristics (N = 34)** |  |
| --- | --- |
| **Characteristic** | **Value** |
| Age (years), mean ± SD | $38.7 \pm 11.2$ |
| Female sex, n (%) | 21 (61.8%) |
| BMI ($\text{kg}/\text{m}^2$), mean ± SD | $25.4 \pm 4.8$ |
| Stable SSRI/SNRI use, n (%) | 14 (41.2%) |
| HAM-A | $28.7 \pm 4.9$ |
| GAD-7 | $16.2 \pm 3.8$ |
| BAI | $27.4 \pm 8.1$ |
| PSS-10 | $25.1 \pm 5.7$ |
| PHQ-9 | $12.8 \pm 5.1$ |
| PSQI | $10.2 \pm 3.6$ |
| RMSSD (ms) | $28.4 \pm 11.7$ |

At baseline, participants demonstrated clinically meaningful moderate-to-severe anxiety severity. Mean baseline HAM-A score was 28.7 ± 4.9, mean GAD-7 score was 16.2 ± 3.8, and mean BAI score was 27.4 ± 8.1. Mean baseline PSS-10 score was 25.1 ± 5.7, mean PHQ-9 score was 12.8 ± 5.1, and mean PSQI score was 10.2 ± 3.6, indicating substantial perceived stress, moderate depressive symptom burden, and impaired sleep quality across the cohort. Stable SSRI/SNRI use was reported in 14 participants (41.2%), Table 1.

### 3.2. Clinician-rated anxiety severity

For the primary clinical outcome, the overall effect of time on HAM-A was significant (F[4, 126] = 7.86, p < 0.001). Mean HAM-A scores decreased from 28.7 ± 4.9 at baseline to 26.1 ± 5.4 at Week 2, 21.8 ± 6.2 at Week 4, 21.6 ± 6.4 at two-week follow-up, and 22.5 ± 6.8 at one-month follow-up. The Week 2 reduction was clinically directional but did not reach statistical significance after correction (estimated mean difference [EMD] = −2.6, 95% CI −5.9 to 0.7, adjusted p = 0.126). In contrast, HAM-A reduction was significant at Week 4 (EMD = −6.9, 95% CI −10.4 to −3.4, adjusted p = 0.001), at two-week follow-up (EMD = −7.1, 95% CI −10.8 to −3.4, adjusted p = 0.001), and at one-month follow-up (EMD = −6.2, 95% CI −10.0 to −2.4, adjusted p = 0.006). The Week 4 change corresponded to a moderate-to-large within-subject effect size (standardized mean change = 0.72), indicating clinically relevant but not complete reduction in clinician-rated anxiety severity by the end of stimulation (Table 2, Table 3).

**Table 2.** Longitudinal Psychological and Physiological Outcomes.

| Outcome | Baseline | Week 2 | Week 4 | FU Week 6 | FU Week 8 |
| --- | --- | --- | --- | --- | --- |
| HAM-A | 28.7 ± 4.9 | 26.1 ± 5.4 | 21.8 ± 6.2 | 21.6 ± 6.4 | 22.5 ± 6.8 |
| GAD-7 | 16.2 ± 3.8 | 15.4 ± 4.1 | 14.2 ± 4.6 | 14.0 ± 4.7 | 14.6 ± 4.9 |
| BAI | 27.4 ± 8.1 | 25.1 ± 8.5 | 21.4 ± 8.0 | 21.1 ± 8.3 | 22.0 ± 8.6 |
| PSS-10 | 25.1 ± 5.7 | 24.5 ± 5.9 | 23.5 ± 6.2 | 23.3 ± 6.4 | 23.9 ± 6.5 |
| PHQ-9 | 12.8 ± 5.1 | 12.3 ± 5.2 | 11.5 ± 5.2 | 11.4 ± 5.3 | 11.8 ± 5.5 |
| PSQI | 10.2 ± 3.6 | 9.9 ± 3.7 | 9.3 ± 3.9 | 9.2 ± 4.0 | 9.5 ± 4.1 |
| RMSSD (ms) | 28.4 ± 11.7 | 31.2 ± 12.4 | 35.1 ± 13.8 | — | — |

**Table 3.** Statistical Summary of Psychological and Physiological Outcomes.

| Outcome | F-statistic | Week 4 EMD (95% CI) | $p$-value | Effect Size |
| --- | --- | --- | --- | --- |
| HAM-A | 7.86 | -6.9 (-10.4, -3.4) | 0.001 | 0.72 |
| GAD-7 | 2.18 | -2.0 (-4.3, 0.3) | 0.089 | — |
| BAI | 4.92 | -6.0 (-10.1, -1.9) | 0.012 | — |
| PSS-10 | 1.21 | -1.6 (-4.4, 1.2) | 0.271 | — |
| PHQ-9 | 1.08 | -1.3 (-3.4, 0.8) | 0.224 | — |
| PSQI | 1.34 | -0.9 (-2.4, 0.6) | 0.238 | — |
| RMSSD (ms) | 4.37 | 6.7 ms (2.1, 11.3) | 0.009 | 0.54 |

### 3.3. Response, remission, and relapse after stimulation discontinuation

At Week 4, 12 participants (35.3%; 95% CI 19.7–53.5%) met predefined clinical response criteria, defined as at least 50% reduction in HAM-A score from baseline. Remission criteria, defined as HAM-A ≤7, were achieved by 3 participants (8.8%; 95% CI 1.9–23.7%). At two-week follow-up, 11 participants (32.4%; 95% CI 17.4–50.5%) remained responders and 2 participants (5.9%; 95% CI 0.7–19.7%) remained in remission. At one-month follow-up, 9 participants (26.5%; 95% CI 12.9–44.4%) remained responders and 2 participants (5.9%; 95% CI 0.7–19.7%) remained in remission.

Relapse was defined exploratorily as loss of Week 4 response status during the stimulation-free follow-up period. Among the 12 Week 4 responders, one participant no longer met response criteria at two-week follow-up and three additional participants had lost response status by one-month follow-up. Thus, relapse among Week 4 responders was 8.3% at two-week follow-up and 33.3% by one-month follow-up. The mean HAM-A trajectory showed a small increase from two-week follow-up to one-month follow-up (21.6 ± 6.4 to 22.5 ± 6.8), suggesting partial maintenance of benefit with mild early rebound after stimulation discontinuation rather than continued progressive improvement.

### 3.4. Secondary self-reported psychological outcomes

Self-reported anxiety outcomes showed a less uniform pattern than clinician-rated HAM-A. The overall time effect for GAD-7 was modest and did not remain robust after correction (F[4, 124] = 2.18, p = 0.075). Mean GAD-7 scores decreased from 16.2 ± 3.8 at baseline to 15.4 ± 4.1 at Week 2, 14.2 ± 4.6 at Week 4, 14.0 ± 4.7 at two-week follow-up, and 14.6 ± 4.9 at one-month follow-up. The Week 4 contrast was borderline and should be interpreted cautiously (EMD = −2.0, 95% CI −4.3 to 0.3, adjusted p = 0.089), while the two-week follow-up and one-month follow-up contrasts were not statistically significant after correction (two-week follow-up: EMD = −2.2, 95% CI −4.8 to 0.4, adjusted p = 0.081; one-month follow-up: EMD = −1.6, 95% CI −4.3 to 1.1, adjusted p = 0.242). Thus, self-reported generalized anxiety improved numerically, but the effect was smaller and less robust than the clinician-rated HAM-A change, Table 2, Table 3, Figure 2.A.

**Figure 2.**
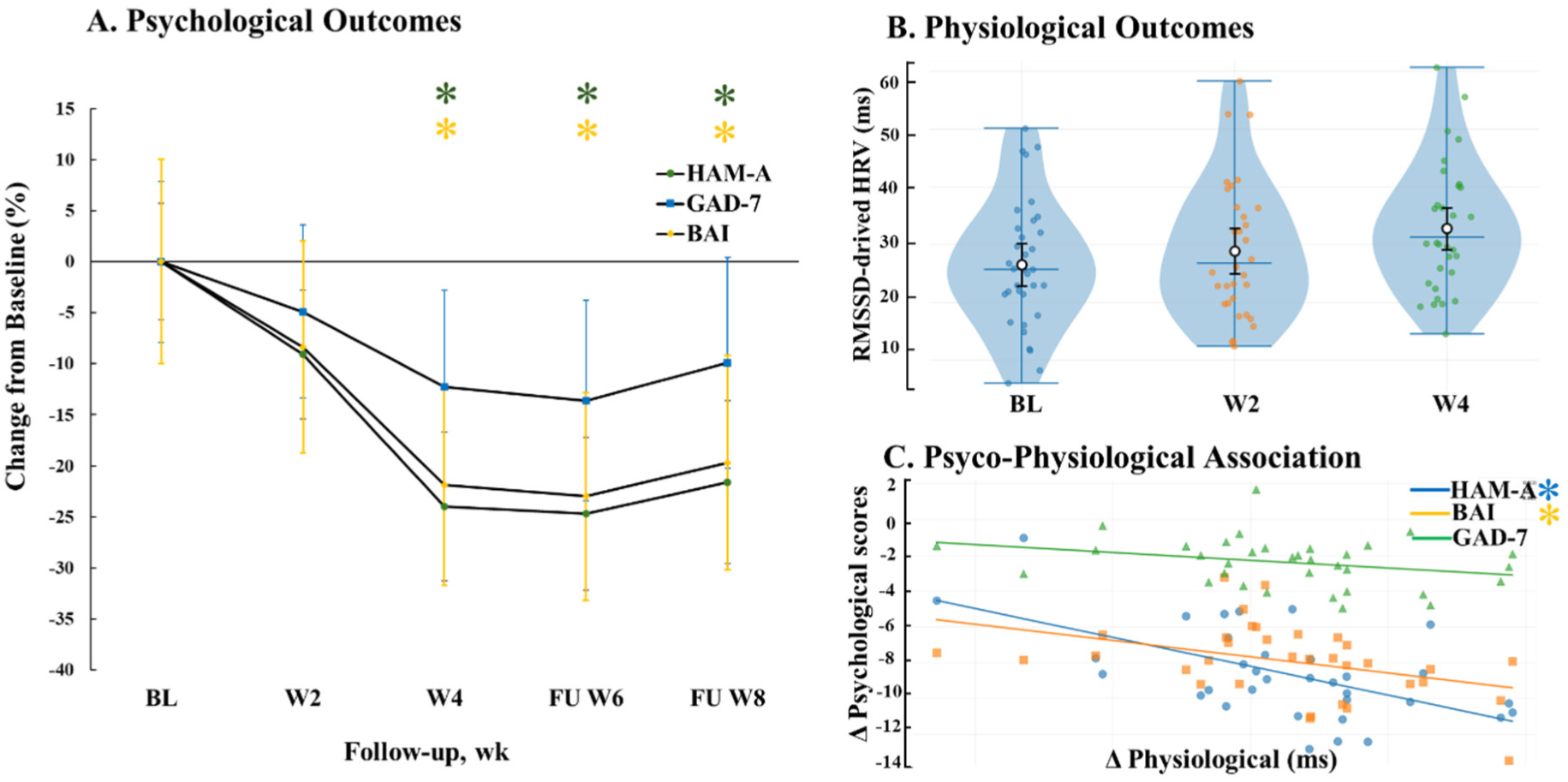
Clinical and physiological outcomes during and after taVNS treatment. A) Longitudinal changes in anxiety measures (HAM-A, GAD-7, and BAI) are shown as percentage change from baseline with 95% confidence intervals. B) RMSSD-derived HRV increased across the intervention period, suggesting enhanced parasympathetic activity. C) Exploratory regression analyses indicate that greater increases in RMSSD were associated with larger improvements in HAM-A and BAI scores, but not with GAD-7. Error bars represent 95% confidence intervals. *p < 0.05. BL: baseline, W: week after the start of intervention, FU: follow up.

BAI scores showed a clearer reduction than GAD-7, consistent with partial improvement in somatic and physiological anxiety symptoms. The overall time effect for BAI was significant (F[4, 123] = 4.92, p = 0.001). Mean BAI scores decreased from 27.4 ± 8.1 at baseline to 25.1 ± 8.5 at

Week 2, 21.4 ± 8.0 at Week 4, 21.1 ± 8.3 at two-week follow-up, and 22.0 ± 8.6 at one-month follow-up. Compared with baseline, the Week 2 change was not significant (EMD = −2.3, 95% CI −6.4 to 1.8, adjusted p = 0.276), whereas reductions were significant at Week 4 (EMD = −6.0, 95% CI −10.1 to −1.9, adjusted p = 0.012) and two-week follow-up (EMD = −6.3, 95% CI −10.7 to −1.9, adjusted p = 0.010). The one-month follow-up effect was attenuated but remained borderline after correction (EMD = −5.4, 95% CI −9.9 to −0.9, adjusted p = 0.047), Table 2, Table 3, Figure 2.A.

Changes in perceived stress, depressive symptoms, and subjective sleep quality were small and did not reach statistical significance after correction. PSS-10 scores decreased from 25.1 ± 5.7 at baseline to 24.5 ± 5.9 at Week 2, 23.5 ± 6.2 at Week 4, 23.3 ± 6.4 at two-week follow-up, and 23.9 ± 6.5 at one-month follow-up. The overall time effect was not significant (F[4, 122] = 1.21, p = 0.310), and the Week 4 contrast remained non-significant (EMD = −1.6, 95% CI −4.4 to 1.2, adjusted p = 0.271). PHQ-9 scores decreased from 12.8 ± 5.1 at baseline to 12.3 ± 5.2 at Week 2, 11.5 ± 5.2 at Week 4, 11.4 ± 5.3 at two-week follow-up, and 11.8 ± 5.5 at one-month follow-up, without a significant overall time effect (F[4, 124] = 1.08, p = 0.371; Week 4 EMD = −1.3, 95% CI −3.4 to 0.8, adjusted p = 0.224). PSQI scores decreased from 10.2 ± 3.6 at baseline to 9.9 ± 3.7 at Week 2, 9.3 ± 3.9 at Week 4, 9.2 ± 4.0 at two-week follow-up, and 9.5 ± 4.1 at one-month follow-up; however, the overall time effect was not statistically significant (F[4, 121] = 1.34, p = 0.260; Week 4 EMD = −0.9, 95% CI −2.4 to 0.6, adjusted p = 0.238), Table 2, Table 3, Figure 2.A.

Overall, the psychological outcome pattern was not uniformly positive across all measures. Clinician-rated anxiety severity and somatic anxiety symptoms showed the clearest improvement, with effects emerging primarily by Week 4 and partially persisting through one-month follow-up. Self-reported generalized anxiety showed only numerical and borderline improvement, while perceived stress, depressive symptoms, and subjective sleep quality showed non-significant numerical changes.

### 3.5. Physiological Outcomes

RMSSD-derived HRV demonstrated a significant change across the intervention period (F[2, 64] = 4.37, p = 0.017). Mean RMSSD increased from **28.4 ± 11.7 ms** during the baseline monitoring window to **31.2 ± 12.4 ms** at the mid-intervention assessment and **35.1 ± 13.8 ms** following completion of the Week 4 stimulation period. Compared with baseline, the increase observed at the mid-intervention window did not reach statistical significance after correction for multiple comparisons (EMD = 2.8 ms, 95% CI −0.8 to 6.4, adjusted p = 0.118). In contrast, RMSSD was significantly higher at the end-of-intervention assessment (EMD = 6.7 ms, 95% CI 2.1 to 11.3, adjusted p = 0.009), corresponding to a moderate within-subject effect size (standardized mean change = 0.54), Table 2, Table 3, Figure 2.B.

Exploratory covariate analyses did not identify significant effects of age, sex, baseline anxiety severity, or stable SSRI/SNRI use on the observed RMSSD trajectory (all p > 0.10). The direction of change was consistent with increased parasympathetic cardiac modulation during repeated taVNS exposure.

### 3.6. Clinical–Physiological Associations

Exploratory regression analysis evaluated whether change in RMSSD was associated with clinical improvement from baseline to Week 4. Greater increases in RMSSD were associated with larger reductions in HAM-A score (β = −0.29, 95% CI −0.55 to −0.03, p = 0.031, R² = 0.18). This finding suggests that improvement in parasympathetic cardiac modulation may track, at least partially, with clinical improvement in clinician-rated anxiety severity. A similar association was observed for BAI scores, where larger increases in RMSSD were associated with greater reductions in somatic anxiety symptoms (β = −0.33, 95% CI −0.60 to −0.06, p = 0.019, R² = 0.21). In contrast, change in RMSSD was not significantly associated with change in GAD-7 scores (β = −0.08, 95% CI −0.31 to 0.15, p = 0.48, R² = 0.02). Although exploratory, these findings suggest a closer relationship between autonomic physiological modulation and somatic manifestations of anxiety than with cognitive worry-related symptoms, Figure 2.C.

### 3.7. Safety, Adverse Events, and Adherence

No serious adverse events occurred among the 48 participants who initiated stimulation during the intervention or follow-up periods. Device-related adverse events were predominantly mild and transient. Among stimulation-exposed participants, the most frequently reported events included local skin irritation at the stimulation site (n = 6, 12.5%), transient tingling discomfort (n = 5, 10.4%), mild headache (n = 4, 8.3%), dizziness during stimulation (n = 2, 4.2%), and temporary nausea (n = 1, 2.1%). Four participants (8.3%) discontinued the intervention because of persistent local stimulation discomfort. Although adverse effects were generally mild, persistent local discomfort was the main tolerability-related reason for discontinuation. No cardiovascular complications, syncopal episodes, or clinically significant neurological adverse events were reported.

Home-based taVNS implementation was feasible across the four-week intervention period among participants included in the final analytic sample. Mean stimulation adherence was 79.8% ± 15.6%, with 25 of 34 participants (73.5%) completing at least 70% of prescribed stimulation sessions. Physiological monitoring feasibility was acceptable, with valid ECG recordings obtained in 91.2% of baseline windows, 85.3% of mid-intervention windows, and 82.4% of end-of-intervention windows after signal quality control, Table 4.

**Table 4.** Safety, Adverse Events, and Adherence.

| **Variable** | **Value** |
| --- | --- |
| Participants initiating intervention | 48 |
| Final analytic sample | 34 |
| Mean stimulation adherence (%) | $79.8 \pm 15.6$ |
| Serious adverse events | 0 |
| Local skin irritation | 6 (12.5%) |
| Tingling discomfort | 5 (10.4%) |
| Headache | 4 (8.3%) |
| Dizziness | 2 (4.2%) |
| Nausea | 1 (2.1%) |
| Withdrawal due to discomfort | 4 (8.3%) |

## 4. Discussion

This prospective single-arm feasibility study evaluated the safety, tolerability, preliminary clinical effects, and exploratory autonomic physiological changes associated with home-based taVNS in adults with psychologist-confirmed moderate-to-severe GAD. Participants completed a four-week home-use stimulation protocol followed by two post-intervention follow-up assessments at two weeks and one month after stimulation discontinuation. In parallel, repeated longitudinal ECG-based physiological monitoring was performed to explore autonomic trajectories during treatment. Overall, the findings suggest that repeated home-based taVNS was safe and generally well tolerated, while preliminary clinical improvements were observed primarily in clinician-rated anxiety severity. In addition, exploratory physiological analyses suggested increases in RMSSD-derived HRV during treatment, with greater RMSSD increases associated with larger reductions in clinician-rated anxiety severity. These findings are discussed in detail below.

### 4.1. Clinical Anxiety Outcomes

From a clinician-rated perspective, anxiety severity decreased over the intervention period, with HAM-A scores demonstrating significant reductions by Week 4 and partial maintenance of improvement during the stimulation-free follow-up period. Importantly, anxiety severity did not improve abruptly during the first two weeks but instead showed progressively larger reductions by the end of stimulation, followed by a mild rebound after stimulation discontinuation. Such temporal trajectories are broadly consistent with previous neuromodulation studies reporting cumulative treatment effects that emerge gradually over repeated stimulation sessions rather than immediately after treatment initiation (Hein et al., 2013; Rong et al., 2016; Straube et al., 2015). The observed pattern may therefore be compatible with progressive neurophysiological adaptation occurring during repeated stimulation exposure.

The magnitude and direction of HAM-A improvement are further consistent with emerging evidence suggesting anxiolytic effects of vagus nerve stimulation across several neurological and psychiatric populations (Gurel et al., 2020; Szeska et al., 2025; Wang et al., 2026). However, direct comparisons remain difficult because previous studies differ regarding stimulation targets, stimulation parameters, treatment duration, outcome measures, patient populations, and methodological rigor (Farmer et al., 2021; Keute et al., 2021). Furthermore, because the present study lacked sham control and randomization, the contribution of other factors, including expectancy effects, placebo responses, increased symptom monitoring, therapeutic engagement, and spontaneous symptom fluctuation, cannot be excluded, but should be addressed in future studies.

In contrast, HAM-A and BAI scores, which place greater emphasis on somatic and autonomic manifestations of anxiety, demonstrated clearer improvement than GAD-7 scores, which primarily assess cognitive worry-related symptoms. This divergence may suggest differential responsiveness across anxiety symptom dimensions. One possible explanation is that the stimulation dosage and treatment duration used in the present study were sufficient to influence physiological hyperarousal and somatic symptom burden but insufficient to induce larger changes in cognitive worry-related symptoms. This interpretation is consistent with evidence from other chronic neurological and psychiatric indications showing that repeated stimulation over longer treatment periods may be required before maximal clinical benefits emerge (Jiao et al., 2020; Rong et al., 2016; Straube et al., 2015). Supportively, several longitudinal neuromodulation studies have adopted repeated daily stimulation schedules over weeks to increase cumulative stimulation exposure and promote progressive clinical improvement (Bauer et al., 2016; Hein et al., 2013; Jiao et al., 2020; Zhang et al., 2024). Given the exploratory nature of the present study, these observations should be interpreted cautiously. Nevertheless, they are broadly compatible with current neurobiological models suggesting that vagal stimulation influences autonomic and interoceptive regulatory circuits involving the nucleus tractus solitarius, locus coeruleus, insula, anterior cingulate cortex, and connected prefrontal–limbic networks (Fang et al., 2016; Frangos et al., 2015; Yakunina et al., 2017). Whether longer treatment duration, alternative stimulation schedules, or combination with psychotherapy may produce larger effects on cognitive worry symptoms remains an important question for future investigation.

Finally, we observed limited changes in perceived stress, depressive symptoms, and subjective sleep quality. The four-week intervention period may have been insufficient to induce broader psychological and behavioral adaptations, as discussed above (Hein et al., 2013; Jiao et al., 2020; Rong et al., 2016; Zhang et al., 2024). In addition, baseline depressive symptom severity in the present cohort was moderate rather than severe, potentially limiting the magnitude of measurable improvement. Finally, subjective sleep quality may improve more slowly than autonomic arousal measures because sleep perception is influenced by multiple behavioral, cognitive, circadian, and environmental factors beyond physiological hyperarousal alone (Buysse, 2014; Riemann et al., 2015). Consequently, the absence of significant changes in these secondary outcomes should not necessarily be interpreted as evidence of a lack of physiological or clinical effect.

### 4.2. Physiological Findings

The observed increase in RMSSD-derived HRV across the intervention period is consistent with enhanced parasympathetic cardiac modulation during repeated taVNS exposure. The direction of change aligns with mechanistic expectations from vagal stimulation models and previous autonomic studies reporting reductions in sympathetic activity and modulation of HRV-related indices during non-invasive vagus nerve stimulation (Clancy et al., 2014; Machetanz et al., 2021). This observation is further supported by evidence demonstrating reduced HRV in anxiety disorders (Chalmers et al., 2014; Friedman, 2007; Kemp et al., 2010) while higher HRV has generally been associated with greater autonomic flexibility, adaptive emotion regulation, and stress resilience (Shaffer and Ginsberg, 2017; Thayer et al., 2009; Thayer and Lane, 2000). Nevertheless, HRV findings following taVNS remain heterogeneous across studies, likely reflecting differences in stimulation protocols, participant populations, recording methodologies, and analytical approaches (Keute et al., 2021; Wolf et al., 2021). Consequently, the present findings should be considered preliminary and require confirmation in larger, sham-controlled studies.

In addition, exploratory clinical–physiological association analyses demonstrated that greater increases in RMSSD were associated with larger reductions in clinician-rated anxiety severity (HAM-A) and somatic anxiety symptoms (BAI), whereas no significant association was observed for GAD-7 scores. This pattern, although exploratory, may be informative. Both the HAM-A and BAI place greater emphasis on autonomic and somatic manifestations of anxiety, including cardiovascular symptoms, muscular tension, restlessness, trembling, and physiological hyperarousal, whereas the GAD-7 predominantly assesses excessive worry and other cognitive anxiety symptoms. The stronger association between RMSSD modulation and improvement in HAM-A and BAI therefore suggests a closer relationship between autonomic physiological regulation and somatic symptom improvement than with cognitive worry-related symptom domains.

This interpretation is biologically plausible given that HRV reflects autonomic nervous system regulation and has repeatedly been linked to vagal tone, stress resilience, emotional regulation, and adaptive physiological flexibility (Shaffer and Ginsberg, 2017; Thayer et al., 2009). Furthermore, emerging evidence suggests that autonomic physiological measures may contribute to the characterization of responsiveness to vagus nerve stimulation interventions (Schiweck et al., 2025). However, these findings should be interpreted cautiously as HRV is influenced by numerous behavioral, circadian, respiratory, hormonal, and environmental factors that may affect its stability and interpretation (Laborde et al., 2017; Quintana and Heathers, 2014). Together, the observed increase in RMSSD and its association with improvements in clinician-rated and somatic anxiety symptoms support the potential utility of HRV as a putative marker of treatment-related physiological change during taVNS. Further studies are however required to determine whether HRV may also contribute to patient stratification, treatment monitoring, and personalized stimulation approaches (Wolf et al., 2021).

### 4.3. Feasibility, Safety, and Home-Based Implementation

A home-based neuromodulation approach can only become clinically meaningful if patients are able to integrate the intervention into daily life with acceptable tolerability and adherence. This is particularly relevant in GAD, a chronic and frequently recurrent condition associated with limited access to psychotherapy, incomplete pharmacological response, and challenges in long-term treatment adherence (Bandelow et al., 2017; Cuijpers et al., 2024; Stein Murray and Sareen, 2015). The findings of the present study suggest that repeated home-based taVNS is feasible and generally well tolerated in adults with moderate-to-severe GAD.

The safety and tolerability profile observed in the current study was broadly consistent with previous taVNS literature (Farmer et al., 2021; Kim et al., 2022; Redgrave et al., 2018). The most commonly reported adverse effects included local skin irritation, transient tingling discomfort, mild headache, and occasional dizziness, all of which have been repeatedly described in prior non-invasive auricular stimulation studies (Burger et al., 2020; Gerges et al., 2024). Importantly, no cardiovascular complications, syncopal episodes, serious adverse events, or clinically significant neurological adverse events were observed. Although local stimulation discomfort contributed to some participant withdrawals, thirty-four participants completed the intervention and follow-up assessments, supporting the practicality of repeated home-based taVNS administration. Future studies should continue to optimize stimulation comfort and user experience while evaluating longer treatment periods and real-world adherence in larger controlled cohorts.

### 4.4. Limitations and Future Directions

Several limitations should be considered when interpreting the present findings. First, the study employed a single-arm observational design without sham control or randomization. Consequently, improvements in anxiety symptoms cannot be attributed exclusively to taVNS, as other factors including expectancy effects, increased symptom monitoring, regression to the mean, and natural symptom fluctuations may have contributed to the observed changes (Farmer et al., 2021; Keute et al., 2021). Second, the sample size was modest and primarily intended to evaluate feasibility, safety, and preliminary clinical signals. The study was therefore not powered to identify clinical subgroups, establish predictors of response, or validate physiological biomarkers.

Third, physiological characterization was limited to ECG-derived autonomic measures. Although RMSSD represents a putative marker of parasympathetic cardiac modulation (Laborde et al., 2017; Shaffer and Ginsberg, 2017) anxiety-related physiological dysregulation also involves respiratory, electrodermal, endocrine, and sleep-related processes that were not comprehensively assessed. Fourth, both the intervention period and follow-up duration were relatively short considering the chronic and recurrent nature of GAD. Longer treatment and observation periods may be required to determine the durability of clinical benefit and to characterize delayed or cumulative neuromodulatory effects (Hein et al., 2013; Rong et al., 2016). Finally, the present study evaluated a single stimulation protocol and therefore cannot inform the optimal stimulation frequency, intensity, treatment duration, session timing, or dosing schedule for anxiety disorders. Previous work suggests that physiological and clinical responses to taVNS may vary considerably across stimulation parameters and individual patient characteristics (Farmer et al., 2021; Machetanz et al., 2021).

Future studies should therefore prioritize adequately powered randomized sham-controlled trials with longer follow-up periods and multimodal physiological assessment. Integration of autonomic, behavioral, endocrine, and neuroimaging biomarkers may improve mechanistic understanding of treatment response and facilitate the development of personalized stimulation approaches. In addition, systematic evaluation of stimulation parameters, treatment schedules, and patient-specific response profiles may help optimize the clinical application of taVNS for anxiety disorders.

## Conclusion

In conclusion, this prospective single-arm feasibility study suggests that home-based taVNS is feasible and generally well tolerated in adults with moderate-to-severe GAD, with preliminary evidence of improvement in clinician-rated anxiety severity, somatic anxiety symptoms, and exploratory autonomic physiological measures. The findings further support the growing interest in non-invasive neuromodulation approaches targeting autonomic and prefrontal–limbic regulatory systems in anxiety disorders. However, because of the observational single-arm design, the results should be interpreted cautiously and primarily as feasibility-oriented. Larger sham-controlled randomized studies are required to determine clinical efficacy, mechanistic specificity, and long-term therapeutic relevance.

## Authorship contribution statement

**Mohsen Mosayebi-Samani:** Conceptualization, Methodology, Formal analysis, Writing – original draft, Visualization, Writing – review & editing. **Erfan Zahirmardi**: Investigation, Data curation, Writing – review & editing. **Saman Hedayat Fard**: Investigation, Data curation, Validation, Writing – review & editing. **Serjhik Azerians**: Supervision, Methodology, Validation, Writing – review & editing.

## Data Availability

All data produced in the present study are available upon reasonable request to the authors.

